# Staff-Pupil SARS-CoV-2 Infection Pathways in Schools: A Population Level Linked Data Approach

**DOI:** 10.1101/2021.02.04.21251087

**Authors:** Daniel A Thompson, Hoda Abbasizanjani, Richard Fry, Emily Marchant, Lucy Griffiths, Ashley Akbari, Joe Hollinghurst, Laura North, Jane Lyons, Fatemeh Torabi, Gareth Davies, Mike B Gravenor, Ronan Lyons

**Affiliations:** Swansea University Medical School, Swansea University, Swansea, UK

## Abstract

**Background:** Better understanding of the role that children and school staff play in the transmission of SARS-CoV-2 is essential to guide policy development on controlling infection whilst minimising disruption to children’s education and wellbeing.

**Methods:** Our national e-cohort (n=500,779) study used anonymised linked data for pupils, staff and associated households linked via educational settings. We estimated the risk of testing positive for SARS-CoV-2 infection for staff and pupils over the period August - December 2020, dependent on measures of recent exposure to known cases linked to their educational settings.

**Results:** The total number of cases in a school was not associated with a subsequent increase in the risk of testing positive (Staff OR per case 0.92, 95%CI 0.85, 1.00; Pupils OR per case 0.98, 95%CI 0.93, 1.02). Amongst pupils, the number of recent cases within the same year group was significantly associated with subsequent increased risk of testing positive (OR per case 1.12, 95%CI 1.08 – 1.15). These effects were adjusted for a range of demographic covariates, and in particular any known cases within the same household, which had the strongest association with testing positive (Staff OR 39.86, 95%CI 35.01, 45.38, pupil OR 9.39, 95%CI 8.94 – 9.88).

**Conclusions:** In a national school cohort, the odds of staff testing positive for SARS-CoV-2 infection were not significantly increased in the 14-day period after case detection in the school. However, pupils were found to be at increased risk, following cases appearing within their own year group, where most of their contacts occur. Strong mitigation measures over the whole of the study period may have reduced wider spread within the school environment.

What is known

- Evidence of the role schools play in the transmission of SARS-CoV-2 is limited
- Higher positivity rates are observed in school staff compared to pupils
- Lack of evidence on transmission pathways transmission into and within schools

What this study adds

- First UK national level study of transmission between pupils and staff in a school environment during the SARS-CoV-2 pandemic.
- Schools opening September-December 2020 was not associated with an increased subsequent risk of testing positive in staff
- Pupils were found to be at increased risk of testing positive, following cases appearing within their own year group

## Introduction

The role schools play in the transmission of severe acute respiratory syndrome coronavirus 2 (SARS-CoV-2) requires further robust evidence. There is ongoing debate regarding closures and related concerns of the negative impacts and widening inequalities in children’s health, wellbeing, educational attainment, as well as on family income and the overall economy. Since the World Health Organization declared the SARS-CoV-2 outbreak a global pandemic on March 11th 2020 (1), education for children and young people has varied from online, in-person and hybrid learning, with wide variance of measures implemented for different groups, within school settings and between countries (2).

Current evidence suggests that younger children are less susceptible to infection (3) and have considerably milder disease compared to adults (4). SARS-CoV-2 positivity rate within the school setting has been low (3,5) and higher positivity rates are observed in school staff compared to pupils (5). In the UK, enhanced surveillance was undertaken following the reopening of schools during the summer half-term 2020, confirming that whilst overall risk of infection was low among pupils and staff, there was a higher risk of SARS-CoV-2 infection among staff and staff-staff transmission was most common (6).

Emerging research from the UK ONS COVID-19 Infection Survey (CIS) and Schools Infection Survey (SIS)(7,8) report increased transmission amongst school staff and school-aged children, particularly aged 12 and above (secondary school age) towards the end of 2020, against a background of high community prevalence. However, the evidence base is still limited and does not cover the dynamics of transmission and infection from households to schools, and within the school setting.

This study contributes to this body of evidence through analyses of population-level data held within the Secure Anonymised Information Linkage (SAIL) Databank (9–11). By linking data on all staff, pupils and associated household contacts in Wales, we aimed to improve understanding of likely transmission pathways into and through educational settings. We assessed the likelihood of test positivity in pupils and staff in relation to other recent cases in linked pupils, staff or their households.

## Methods

### e-Cohort Creation

We created an e-cohort of school children (ages 4-17), school staff, and linked household members for both children and staff (Figure 1). The e-cohort was created using anonymised linked data held within the SAIL Databank at Swansea University (9–11). Data are anonymised at an individual and household level (12,13). Our primary health data cohort was the Welsh COVID-19 e-cohort (14) which consists of all people alive and known to the NHS in Wales on or after the 1st January 2020. To this core we linked the School Workforce Annual Census (SWAC) which details all individuals who work in a publicly funded school (15) covering 1498 out of 1502 schools in Wales; and the Pupil Level Annual School Census (PLASC)(16) which includes annual returns on 1480 out of 1502 schools. Finally, we linked COVID-19 antigen testing data to the cohort. This data combined pillar 1 and pillar 2 data collected by Public Health Wales (PHW) (17). Pillar 1 is swab testing in PHW labs and NHS hospitals for those with a clinical need, and health and care workers; and Pillar 2 is swab testing for the wider population, as set out in government guidance. These linkages are summarised in Figure 1.

**Figure 1.**
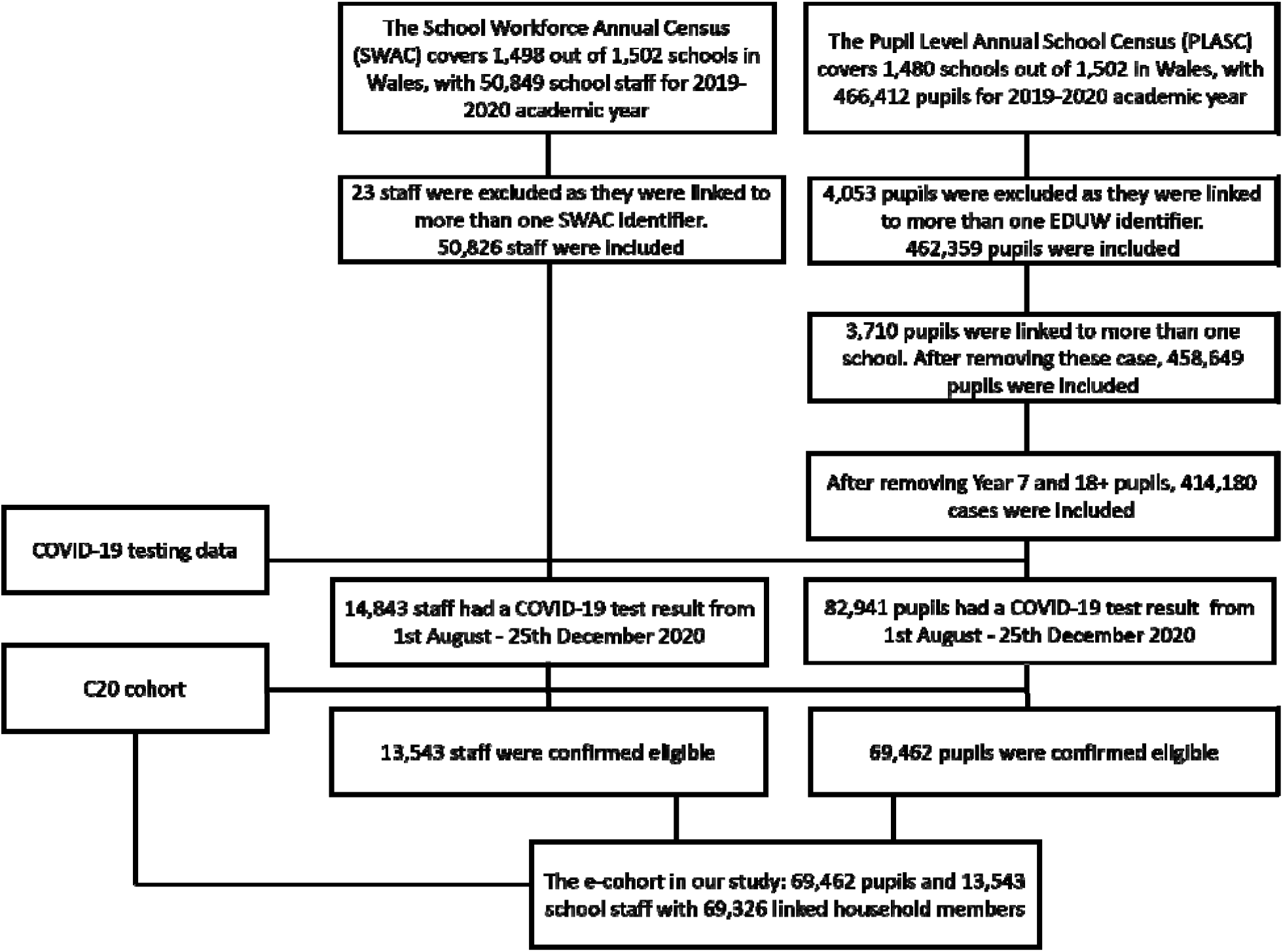
Health and administrative education data linkages linked via SALF. Missing variables of staff and pupils before being confirmed eligible are reported in Table S3.

Our e-cohort study used pupils, staff and linked household members in Wales linked via educational settings using a School Anonymised Linking Field (SALF). We followed participants from 2020-08-01 to 2020-12-25. Our educational setting data is recent up to the end of the academic year 2019-2020. Therefore, we removed pupils who: finished primary school (year 6) in the school year 2019/20; finished secondary school (year 11) in the school year 2019/20 from counts of outcomes within an educational setting and the statistical models, because it is not possible to confirm their linked education setting over the period. Staff members contracted to multiple schools (i.e. peripatetic teachers) were also removed because it was not possible to determine durations within each school.

### Patient Public Involvement

All proposals to use anonymised data in SAIL are scrutinised by an independent Information Governance Review Panel (IGRP) that includes members of the public prior to the commencement of the research.

### Statistical Modelling

Our outcome was the probability of testing positive, following a pillar 1 or pillar 2 test. The outcome was determined by the number of school-linked positive cases in the preceding 14-day period, prior to the collection date of the outcome’s specimen (date-of-interest). Exposure measures investigated were: 1) total number of cases within the linked school, 2) total number of cases within the linked household, 3) total number of cases in any households linked to the school, 4) total number of cases within the same year group (pupils only), which represents the pupil population in which the vast majority of contacts for an individual pupil would occur.

We used binary logistic regression to determine the odds ratios (ORs) for a positive outcome after a SARS-CoV-2 test. We first combined both staff and pupils test results to determine general associations (Model M1), with a categorical variable indicating whether an individual was a staff or a pupil member at the linked school. We then stratified by staff (M2) and pupil outcomes (M3). Individuals with any missing covariate data were removed. As additional covariates, we included age, sex, rurality (18), school type and number of staff and pupils in the same school.

## Results

### Cohort Characteristics

The study was based on 500,779 pupils and staff attending schools in Wales. Details of numbers, school categories, tests and percentage positive are shown in Table 1.

**Table 1:** Cohort Summary

|  | Individuals<br>(% of total) | Tested<br>(% of population) | Positive Results<br>(% of population) |
| --- | --- | --- | --- |
| Staff | 50,826 (10.1%) | 15,676 (30.8%) | 3,256 (6.4%) |
| Pupils (16+) | 42,213 (8.4%) | 4,608 (10.9%) | 408 (1.0%) |
| Pupils (12-16) | 166,031<br>(33.2%) | 21,440 (12.9%) | 903 (0.5%) |
| Pupils (<11 incl.<br>nursery) | 241,709<br>(48.3%) | 31,544 (13.1%) | 715 (0.3%) |
| **Total** | **500,779** | **73,268 (14.6%)** | **5,282 (1.1%)** |

### Potential Routes of Transmission

Table 2 summarises the different settings in which potential exposure to the SARS-CoV-2 virus can be identified, based on a time window of 14-days preceding a positive test. The large majority of pupils and staff had a recorded exposure in either their household or school. There were recent potential exposures at school for 75% of positive staff, with 58% having school-but-not-household exposure. For pupils, 85% had recent school cases, with 44% having school-but-not-household.

**Table 2:** Distribution of known potential exposure to infection by setting for staff and pupils

| Exposure for staff and pupils in the 14-day preceding window of their first SARS-CoV-2 positive test from 2020-08-01 to 2020-12-25 |  |  |  |  |  |  |
| --- | --- | --- | --- | --- | --- | --- |
| Setting | Staff |  |  | Pupils |  |  |
|  | *n* | *% of positive cases* | *% of total cohort* | *n* | *% of positive cases* | *% of total cohort* |
| School-only exposure | 1766 | 58.46% | 3.47% | 3352 | 43.59% | 0.74% |
| Household-only exposure | 240 | 7.94% | 0.47% | 634 | 8.25% | 0.14% |
| Both school and household exposure | 500 | 16.55% | 0.98% | 3058 | 39.78% | 0.68% |
| Neither House nor School | 515 | 17.05% | 1.01% | 644 | 8.38% | 0.14% |

### Effect of school exposure on risk of positive test

In unadjusted analyses (supplementary information, Tables S1 and S2), we found significantly increased risk of testing positive across all outcomes, following known cases in linked schools and households. However, after adjusting for age, sex, rurality, school type, household case exposure, and numbers of staff/pupils in school/household, we found that total numbers of cases in the preceding 14 days in the school was associated with a lower risk of testing positive (Staff OR 0.93, 95%CI 0.89, 0.97; pupils OR 0.97, 95%CI 0.95 – 0.98; table 3 M1).

**Table 3:** Logistic Regression Results (M1 Staff and Pupils; M2 Stratified by Staff; M3 Stratified by Pupils). Adjustments for age, sex, residential settlement type, number of pupils and staff within the linked school, and number of people within linked household are included in the models. Odds ratios calculated per individual case of known exposure.

| Exposure variable<br>(within last 14 days) | M1 Staff and<br>Pupil Outcomes<br>(n = 83,004) | M2 – Staff<br>Outcomes<br>(n = 13,543) | M3 – Pupil<br>Outcomes<br>(n = 69,461) |
| --- | --- | --- | --- |
| Count of cases within own household | 11.81***<br>(11.02 – 12.15) | 39.86***<br>(35.01 – 45.38) | 9.39***<br>(8.94 – 9.88) |
| Count of staff member cases within the linked school | 0.93***<br>(0.89 – 0.97) | 0.92'<br>(0.85 – 1.00) | 0.97<br>(0.91– 1.01) |
| Count of pupil cases within the linked school (non-year for M3) | 0.97***<br>(0.95 – 0.98) | 0.98<br>(0.93 – 1.02) | 0.92***<br>(0.89 – 0.94) |
| Count of pupil cases in the linked school within the same year group | - | - | 1.12***<br>(1.08 – 1.15) |
| Count of cases in staff member's homes linked to the school. | 1.11***<br>(1.07 – 1.15) | 1.09**<br>(1.02 – 1.17) | 1.17***<br>(1.12 – 1.22) |
| Count of cases in pupils' homes linked to the school. | 1.07***<br>(1.06 – 1.09) | 1.04*<br>(1.01 – 1.07) | 1.08***<br>(1.06 - 1.10) |

Signif. codes: 0 '\*\*\*' 0.001 '\*\*' 0.01 '\*' 0.05 '.' 0.1 ' ' 1

Unsurprisingly, by far the strongest signal in the data (for both staff and pupils) related to exposure to known cases in the household (table 3, M1-3). We also found a significant association with the wider bubble of cases in any household linked to the school (table 3, M1-3).

When stratifying by staff test results, and after adjusting for covariates (including household cases), the total number of cases occurring in a linked school setting was again associated with slightly lower odds of a positive SARS-CoV-2 outcome (OR 0.92, 95%CI 0.85, 1.00 and 0.98, 95%CI 0.93, 1.02 for exposure to staff and pupil cases respectively). Staff members in primary and special schools had a higher odds of a SARS-CoV-2 positive test compared with middle and secondary schools, and staff had higher odds of a positive outcome compared to the reference level of pupils (OR 2.99, 95%CI 1.67-5.37, p value <0 .001).

When stratifying by pupils, and adjusting for covariates (including household cases), the total number of cases in the school was not associated with increased risk of test positivity (Table 3). However, in contrast, the number of cases in *pupils within the same year* group was significantly associated with testing positive (OR 1.12, 95%CI 1.08-1.15).

## Discussion

### Summary of main findings

Our results show that the total number of SARS-CoV-2 positive staff and pupils within a school following the re-opening in Wales in September 2020 was not associated with an increased subsequent risk of testing positive in staff or pupils. By including likely household exposure and number of cases in all households linked to the school in the models, we aimed to adjust for one of the primary routes of transmission (own household), and also a proxy measure of community prevalence, which increased considerably over the period. The lack of association at the school level sheds light on the effectiveness of reducing transmission within the school environment, and also on the policy of isolation following exposure (19). Wales adopted an aggressive policy of school year group (secondary), school class (primary) and large bubble closures following the detection of cases, even when prevalence was low. Notably, the numbers of pupils in schools declined dramatically during the period of highest prevalence in December. Average pupil attendance was approximately 85% until the end of November, but dropped to 70% by the 7 ^th^ December and 33% by the 14 ^th^.

Nevertheless, our results also demonstrate increased odds of a SARS-CoV-2 positive outcome in pupils dependent on the number of cases found in the same year group. As this represents by far the majority of contacts for all schoolchildren, the results are consistent with pupil-pupil transmission. We estimated a 12% increase in the odds of testing positive, for case in the year group in the preceding exposure window (75% increase for 5 cases). It is notable that this signal can be detected after adjustment for household exposure, some measures of community prevalence, and especially amidst a background of active isolation measures.

Unsurprisingly, SARS-CoV-2 infections within an individual’s household posed a highly significant risk of subsequent infection in school staff and pupils. In addition, the number of SARS-CoV-2 positive outcomes within any households linked to the school also suggest increased odds of a SARS-CoV-2 positive outcome in staff and pupils. This may reflect a direct effect of contacts occurring around the school environment, or also be a general marker of community prevalence. We noted that very few cases were recorded who did not have a link to a known case in either the home or school environment. Furthermore, a large majority of both staff and pupils were potentially exposed to school cases, while having no known household exposure.

### Comparison with previous work

Public health responses, and decisions on school closures, are informed by the best available evidence. This is rapidly evolving and a number of reviews have been published recently (2,20) some of which include primary studies on transmission during the first wave, and others which look at the situation across 2020. A recent review highlighted the large heterogeneity amongst studies investigating the impact of school closures and reopening schools on transmission (21).

There is consistent evidence that children aged below 10 to 14 years have lower susceptibility to SARS-CoV-2 infection than adults (3,20) and that children play a limited role in overall transmission rates. However, there remains few high-quality studies that disentangle potential transmission routes between households and schools, and transmission of SARS-CoV-2 within the school setting between pupils and school staff (21). Our study contributes to this gap in the evidence base, and demonstrates that transmission risks in schools exist, but likely are at much lower than in households as long as other mitigation measures are in place.

The balance of evidence thus far indicates low overall positivity rates in the school environment (5). A low overall risk of infection among staff and pupils within educational settings have been observed in countries that remained open for face-to-face teaching during the first wave in Spring 2020 in Australia (22) and Sweden (4). These studies concluded that the attendance of children and school staff within educational settings maintaining physical distancing and hygiene measures did not contribute substantially to overall infection rates. Following national school closures and the reopening of schools in the summer term of 2020, evidence from Israel (23) suggested that schools reopening had a limited effect on SARS-CoV-2 infection rate in children and adults, and national surveillance in England found low overall risk of infection among staff and pupils in educational settings, although staff-staff transmission was most common (6). Our study extends this evidence base by examining if transmission varied between and within year groups. Our results show pupil-pupil transmission within a year group may occur before cases are identified, but current measures including rapid isolation and implementing physical distancing such as segregated year groups may be effective in reducing the scale of this, and containing subsequent transmission within the school.

In a similar time period to the current study (August to December 2020), evidence from Canada (24) examined secondary transmission of SARS-CoV-2 and reported no instances of child-to-adult transmission during in-person teaching. Whilst findings from the current study reflect that of largely symptomatic testing of pupils and staff, contact tracing during this period of all children (symptomatic and asymptomatic) under 14 years exposed to a confirmed case and tested during the following 14 day isolation period showed minimal pupil-pupil and pupil-staff transmission in primary schools situated within two counties in Norway with high community incidence (25). Consistent with other studies is our finding of higher positivity rates among school staff compared to pupils (5,6,22) and may reflect the higher population-based rates observed in adults.

### Study strengths and limitations

Our study included the entire staff and pupil records in Wales, in publicly funded schools, and hence avoids some selection biases, other than through the privately educated sector, which is very small in Wales (75 private schools). The sample size of tests, and numbers of infections was substantial. A key strength is the fine scale of data linkage, which allowed us to link household and school events, which has not been a feature in previous reports. Adjusting for likely transmission in the home and through extended school bubbles is important in clarifying effect sizes for likely transmission in the school and community setting.

Among the weaknesses of our study design is that testing for cases has been very largely based on testing those who are symptomatic, and most staff and pupils have not been tested. Hence, our results are based on detected cases and not all infections. The school links are generated from 2019 data. Some pupils will have left or moved school during the summer holidays which could introduce biases. To mitigate against this, we excluded all children aged 11 or 16+ in the 2019 data as these will have moved from primary to secondary schools or have left school. We cannot exclude that there will be some mismatches with linking children to schools they no longer attend.

Measures to reduce transmission in the school environment, although advised at a national government level, will likely have varied subtly across schools in Wales dependent on setting, numbers of staff available and personal behaviours of children, staff and parents (e.g. mask wearing and congregating at school opening and closing times). We are unable to capture these variations in routine data which may explain some of the differences observed and we have also not examined new variants of SARS-CoV-2. Finally, we are currently unable to account for days when pupils may not have been present in school, which may have resulted in different exposures for a small number of cases.

### Implications

National school closures are a topic of ongoing debate regarding the risks and benefits between potential transmission within the school setting, balanced against concerns of the negative impacts and widening inequalities in children’s health, wellbeing and educational attainment, and the broader economic and societal impact. Findings from this study suggest that pupil to pupil SARS-CoV-2 transmission is likely but the absolute effects on the wider school population and staff can be minimised through the implementation of current mitigation measures, albeit measures that have been strict. Approximately 15% of the pupil population was absent from school over most of the study period, increasing to 70% as the second wave peak approached, with early complete Christmas closure.

This study has examined plausible transmission pathways within a school environment and not the risk of staff or pupils becoming moderately or seriously ill from COVID-19. As there is a paucity of evidence on the effectiveness of the vaccines on the reduction of transmission it is beyond the scope of this paper to assess whether educational staff should be re-prioritised for vaccination. However, as the vaccines are rolled out further urgent work is warranted to examine the effectiveness of vaccines in reducing transmission within educational settings.

## Conclusion

This study has shown that there are significant complexities in understanding the vectors for transmission within schools. Whilst this study has been conducted in Wales it is highly likely that the findings are generalisable to the UK and many parts of the world in temperate climates where schools have around 30 pupils per class and are largely educated indoors. We conclude that there is good evidence that the numbers of cases in pupils is associated with exposure to previous pupil cases within the school year group, consistent with pupil-pupil transmission linked to schools. A wide range of extensive mitigation measures in our study population have likely reduced the potential for further spread within the wider school pupil population and from pupil to staff.

## Supporting information

STROBE Checklist

## Data Availability

All data used in this study are anonymised. Access to the data is available on application to the SAIL Databank (www.saildatabank.com).

## Ethics

The data used in this study are available in the SAIL Databank at Swansea University, Swansea, UK. All proposals to use SAIL data are subject to review by an independent Information Governance Review Panel (IGRP). Before any data can be accessed, approval must be given by the IGRP. The IGRP gives careful consideration to each project to ensure proper and appropriate use of SAIL data. When access has been approved, it is gained through a privacy-protecting safe haven and remote access system referred to as the SAIL Gateway. SAIL has established an application process to be followed by anyone who would like to access data via SAIL https://www.saildatabank.com/application-process.

All research conducted in this study has been completed under the permission and approval of the SAIL independent Information Governance Review Panel (IGRP) project number 0911.

## Acknowledgements

This work uses data provided by patients and collected by the NHS as part of their care and support. We would also like to acknowledge all data providers who make anonymised data available for research.

We wish to acknowledge the collaborative partnership that enabled acquisition and access to the de-identified data, which led to this output. The collaboration was led by the Swansea University Health Data Research UK team under the direction of the Welsh Government Technical Advisory Cell (TAC) and includes the following groups and organisations: the Secure Anonymised Information Linkage (SAIL) Databank, Administrative Data Research (ADR) Wales, NHS Wales Informatics Service (NWIS), Public Health Wales, NHS Shared Services Partnership and the Welsh Ambulance Service Trust (WAST). All research conducted has been completed under the permission and approval of the SAIL independent Information Governance Review Panel (IGRP) project number 0911.

## Author Contributions

DT and HA led the design, analysis and drafting of the paper.

All other authors contributed equally to the design, data acquisition and interpretation of the data and reviewed the manuscript contents.

All authors have approved the final published version.

## References

1. World Health Organization. WHO Director-General’s opening remarks at the media briefing on COVID-19 - 11 March 2020 [Internet]. [cited 2021 Feb 2]. Available from: https://www.who.int/director-general/speeches/detail/who-director-general-s-opening-remarks-at-the-media-briefing-on-covid-1911-march-2020

2. Krishnaratne S, Pfadenhauer LM, Coenen M, Geffert K, Jung-Sievers C, Klinger C, et al. Measures implemented in the school setting to contain the COVID-19 pandemic: a scoping review. Cochrane database Syst Rev. 2020;12:CD013812.

3. Viner RM, Mytton OT, Bonell C, Melendez-Torres GJ, Ward J, Hudson L, et al. Susceptibility to SARS-CoV-2 Infection among Children and Adolescents Compared with Adults: A Systematic Review and Meta-analysis. JAMA Pediatrics. 2020.

4. Ludvigsson JF. Systematic review of COVID-19 in children shows milder cases and a better prognosis than adults. Vol. 109, Acta Paediatrica, International Journal of Paediatrics. 2020.

5. Xu W, Li X, Dozier M, He Y, Kirolos A, Mathews C, et al. What is the evidence for transmission of COVID-19 by children in schools? A living systematic review. J Glob Health. 2020 Dec 1;10(2).

6. Ismail SA, Saliba V, Lopez Bernal J, Ramsay ME, Ladhani SN. SARS-CoV-2 infection and transmission in educational settings: a prospective, cross-sectional analysis of infection clusters and outbreaks in England. Lancet Infect Dis. 2020;

7. Coronavirus (COVID-19) Infection Survey, UK Statistical bulletins - Office for National Statistics [Internet]. [cited 2021 Feb 2]. Available from: https://www.ons.gov.uk/peoplepopulationandcommunity/healthandsocialcare/conditionsanddiseases/bulletins/coronaviruscovid19infectionsurveypilot/previousReleases

8. COVID-19 Schools Infection Survey Round 1, England - Office for National Statistics [Internet]. [cited 2021 Feb 2]. Available from: https://www.ons.gov.uk/peoplepopulationandcommunity/healthandsocialcare/conditionsanddiseases/bulletins/covid19schoolsinfectionsurveyround1england/november2020

9. Lyons RA, Jones KH, John G, Brooks CJ, Verplancke J-P, Ford D V, et al. The SAIL databank: linking multiple health and social care datasets. BMC Med Inform Decis Mak. 2009 Dec;9(1):3.

10. Ford D V, Jones KH, Verplancke J-P, Lyons RA, John G, Brown G, et al. The SAIL Databank: building a national architecture for e-health research and evaluation. BMC Health Serv Res. 2009 Dec;9(1):157.

11. Jones KH, Ford D V, Jones C, Dsilva R, Thompson S, Brooks CJ, et al. A case study of the Secure Anonymous Information Linkage (SAIL) Gateway: a privacy-protecting remote access system for health-related research and evaluation. J Biomed Inform. 2014;50:196–204.

12. Rodgers SE, Lyons RA, Dsilva R, Jones KH, Brooks CJ, Ford D V, et al. Residential Anonymous Linking Fields (RALFs): A novel information infrastructure to study the interaction between the environment and individuals’ health. J Public Health (Bangkok). 2009;31(4):582–8.

13. Johnson RD, Griffiths LJ, Hollinghurst J, Akbari A, Lee A, Thompson DA, et al. Deriving household composition using population-scale Electronic Health Record data – a reproducible methodology.

14. Lyons J, Akbari A, Torabi F, Davies GI, North L, Griffiths R, et al. Understanding and responding to COVID-19 in Wales: Protocol for a privacy-protecting data platform for enhanced epidemiology and evaluation of interventions. BMJ Open. 2020;10(10).

15. Government W. School Workforce Annual Census data: background, quality and methodology information. 2020.

16. Pupil level annual school census (PLASC) | Sub-topic | GOV.WALES [Internet]. [cited 2021 Feb 2]. Available from: https://gov.wales/pupil-level-annual-school-census-plasc

17. Department of Health & Social Care. COVID-19 testing data: methodology note. 2020.

18. Office for National Statistics. 2011 rural/urban classifications. 2016.

19. Schools: coronavirus guidance | GOV.WALES [Internet]. [cited 2021 Feb 3]. Available from: https://gov.wales/schools-coronavirus-guidance#section-52383

20. Li X, Xu W, Dozier M, He Y, Kirolos A, Theodoratou E. The role of children in transmission of SARS-CoV-2: A rapid review. J Glob Health. 2020;10(2).

21. Walsh S, Chowdhury A, Russell S, Braithwaite V, Ward J, Waddington C, et al. Do school closures reduce community transmission of COVID-19? A systematic review of observational studies. medRxiv. 2021;2021.01.02.21249146.

22. Macartney K, Quinn HE, Pillsbury AJ, Koirala A, Deng L, Winkler N, et al. Transmission of SARS-CoV-2 in Australian educational settings: a prospective cohort study. Lancet Child Adolesc Heal. 2020;4(11).

23. Somekh I, Shohat T, Boker LK, Simões EAF, Somekh E. Reopening Schools and the Dynamics of SARS-CoV-2 Infections in Israel: A Nationwide Study. Clin Infect Dis. 2021;

24. Zimmerman KO, Akinboyo IC, Brookhart MA, Boutzoukas AE, McGann K, Smith MJ, et al. Incidence and Secondary Transmission of SARS-CoV-2 Infections in Schools. Pediatrics. 2021;e2020048090.

25. Brandal LT, Ofitserova TS, Meijerink H, Rykkvin R, Lund HM, Hungnes O, et al. Minimal transmission of SARS-CoV-2 from paediatric COVID-19 cases in primary schools, Norway, August to November 2020. Eurosurveillance. 2021;26(1).

