## Supplementary Tables for "Staff-Pupil SARS-CoV-2 Infection Pathways in Schools: A Population Level Linked Data Approach"

Table S1 – Odds ratios of Univariate models for exposure variables. Signif. codes: 0 '***' 0.001 '**' 0.01 '*' 0.05 '’' 0.1 ' ' 1.

| **Exposure variable**  **(within last 14 days)** | **Model UA 1**  (unadjusted)  n = 83,004 | **Model UA 2**  (unadjusted)  n = 83,004 | **Model UA 3**  (unadjusted)  n = 83,004 | **Model UA 4**  (unadjusted)  n = 69,461 | **Model UA 5**  (unadjusted)  n = 69,461 | **Model UA 6**  (unadjusted)  n = 83,004 | **Model UA 7**  (unadjusted)  n = 83,004 |
| --- | --- | --- | --- | --- | --- | --- | --- |
| Count of cases within own household | 8.20***  (8.19 – 8.81) |  |  |  |  |  |  |
| Count of staff member cases within the linked school |  | 1.17***  (1.16 – 1.18) |  |  |  |  |  |
| Count of pupil cases within the linked school |  |  | 1.12***  (1.11 – 1.12) |  |  |  |  |
| Count of pupil cases within the linked school (non-year group) |  |  |  | 1.19***  (1.18 – 1.20) |  |  |  |
| Count of pupil cases in the linked school within the same year group |  |  |  |  | 1.48***  (1.45 – 1.50) |  |  |
| Count of cases in staff member’s homes linked to the school. |  |  |  |  |  | 1.47***  (1.16 – 1.18) |  |
| Count of cases in pupils' homes linked to the school. |  |  |  |  |  |  | 1.09***  (1.08 – 1.09) |

| **Confounder Variable** | **Category** | **MS1**  **n = 83,004** | **MS2**  **n = 83,004** | **MS3**  **n = 83,004** | **MS4**  **n = 83,004** | **MS5**  **n = 83,004** | **MS6**  **n = 83,004** | **MS7**  **n = 83,004** | **MS9**  **n = 83,004** | **MS10 n = 83,004** |
| --- | --- | --- | --- | --- | --- | --- | --- | --- | --- | --- |
| Age Group | 0 - 4 (ref) |  |  |  |  |  |  |  |  |  |
|  | 0 - 9 | 1.62*** (1.47 - 1.78) |  |  |  |  |  |  |  | 1.57*** (1.43 - 1.73) |
|  | 10 – 14 | 2.49*** (2.27 - 2.73) |  |  |  |  |  |  |  | 2.69*** (2.40 - 3.02) |
|  | 15 – 19 | 3.63*** (3.15 - 4.19 |  |  |  |  |  |  |  | 3.84*** (3.25 - 4.53) |
|  | 20 – 24 | 5.16*** (4.20 - 6.34) |  |  |  |  |  |  |  | 2.18** (1.28 - 3.71) |
|  | 25 – 29 | 4.60*** (3.96 - 5.34) |  |  |  |  |  |  |  | 2.16** (1.29 - 3.34) |
|  | 30 – 34 | 4.55*** (3.96 - 5.24) |  |  |  |  |  |  |  | 2.01** (1.21 - 3.08) |
|  | 35 – 39 | 4.41*** (3.82 - 5.24) |  |  |  |  |  |  |  | 1.85* (1.11 - |
|  | 40 – 44 | 5.59*** (4.77 - 6.25) |  |  |  |  |  |  |  | 2.23** (1.34 - 3.71) |
|  | 45 – 49 | 5.58*** (4.89 - 6.39) |  |  |  |  |  |  |  | 2.39*** (1.43 - 3.97) |
|  | 50 – 54 | 5.50*** (4.77 - 6.35) |  |  |  |  |  |  |  | 2.45*** (1.47 - 4.09) |
|  | 55 – 59 | 5.39*** 4.53 - 6.40) |  |  |  |  |  |  |  | 2.55*** (1.52 - 4.29) |
|  | 60 – 64 | 5.52*** (4.22 - 7.21) |  |  |  |  |  |  |  | 2.70*** (1.54 - 4.74) |
|  | 65 – 69 | 2.77* (1.25 - 6.12) |  |  |  |  |  |  |  | 1.47 (0.58 - 3.76) |
|  | 70 – 74 | 4.39** (1.48 - 13.01) |  |  |  |  |  |  |  | 2.35 (0.71 -7.76) |
|  | 75 – 79 | 14.82*** (3.32 - 66.35) |  |  |  |  |  |  |  | 7.96* (1.64 - 38.68) |
| Gender | Male( ref.) |  |  |  |  |  |  |  |  |  |
|  | Female |  | 1.37*** (1.31 - 1.44) |  |  |  |  |  |  | 1.04* (0.99 - 1.09) |
| Residential Settlement Type | Rural town and fringe in sparse setting |  |  | 0.51*** (0.41 - 0.64) |  |  |  |  |  | 0.60*** (0.47 - 0.75) |
|  | Rural village and dispersed |  |  | 0.90' |  |  |  |  |  | 0.89’ (0.79 - 1.01) |
|  | Rural village and dispersed in a sparse setting |  |  | 0.64*** (0.41 - 1.02) |  |  |  |  |  | 0.69*** (0.60 - 0.81) |
|  | Urban city and town |  |  | 1.16** (1.09 - 1.24) |  |  |  |  |  | 1.13*** (1.05 - 1.21) |
|  | Urban city and town in a sparse setting |  |  | 0.70** (0.54 - 0.89) |  |  |  |  |  | 0.79’ (0.61 - 1.01) |
|  | Rural town and fringe (ref.) |  |  |  |  |  |  |  |  |  |
| School Type | Primary (ref.) |  |  |  |  |  |  |  |  |  |
|  | Middle |  |  |  | 1.04 (0.94 - 1.16) |  |  |  |  | 0.60*** (0.53 - 0.69) |
|  | Nursery or PRU |  |  |  | 0.33* (0.14 - 0.82) |  |  |  |  | 0.42’ (0.17 - 1.03) |
|  | Secondary |  |  |  | 1.47*** (1.40 - 1.54) |  |  |  |  | 0.81*** (0.74 - 0.89) |
|  | Special |  |  |  | 1.78*** (1.58 - 2.02) |  |  |  |  | 0.81** (0.69 - 0.95) |
| School relation | Pupil (ref.) |  |  |  |  |  |  |  |  |  |
|  | Staff |  |  |  |  | 2.74*** (2.60 - 2.88) |  |  |  | 2.72*** (1.77 - 4.69) |
| Number of staff within school |  |  |  |  |  |  | 1.01*** (1.01 - 1.01) |  |  | 1.00** (1.00 - 1.00) |
| Number of pupils within the school |  |  |  |  |  |  |  | 1.01*** (1.01 - 1.01) |  | 1.00*** (1.00 - 1.00) |
| Number within household |  |  |  |  |  |  |  |  | 1.26*** (1.24 - 1.28) | 1.35*** (1.33 - 1.38) |

Table S2 – Odds ratios of Univariate models for confounder variables. Signif. codes: 0 '***' 0.001 '**' 0.01 '*' 0.05 '’' 0.1 ' ' 1.

| **Missing variable** | **Total number of individuals with missing variable** |
| --- | --- |
| RALF | 4,778 |
| Residential Settlement Type | 5,587 |
| School Type | 350 |
| Number of staff within the school | 842 |
| Number of pupils within the school | 189 |
| Other (data linkage issue) | 1,508 |

Table S3 – Number of individuals with missing variables before being confirmed eligible for the cohort (reference to Figure 1). Note this is not a count of distinct individuals, multiple persons may have multiple missing variables.
